# On Topological Properties of COVID-19: Predicting and Controling Pandemic Risk with Network Statistics

**DOI:** 10.1101/2020.09.17.20197020

**Authors:** Mike K.P. So, Amanda M.Y. Chu, Agnes Tiwari, Jacky N.L. Chan

## Abstract

The spread of coronavirus disease 2019 (COVID-19) has caused more than 24 million confirmed infected cases and more than 800,000 people died as of 28 August 2020. While it is essential to quantify risk and characterize transmission dynamics in closed populations using Susceptible-Infection-Recovered modeling, the investigation of the effect from worldwide pandemic cannot be neglected. This study proposes a network analysis to assess global pandemic risk by linking 164 countries in pandemic networks, where links between countries were specified by the level of ‘co-movement’ of newly confirmed COVID-19 cases. More countries showing increase in the COVID-19 cases simultaneously will signal the pandemic prevalent over the world. The network density, clustering coefficients, and assortativity in the pandemic networks provide early warning signals of the pandemic in late February 2020. We propose a preparedness pandemic risk score for prediction and a severity risk score for pandemic control. The preparedness risk score contributed by countries in Asia is between 25% to 50% most of the time after February and America contributes close to 50% recently. The high preparedness risk contribution implies the importance of travel restrictions between those countries. The severity risk score of America is greater than 50% after May and even exceeds 75% in July, signifying that the control of COVID-19 is still worrying in America. We can keep track of the pandemic situation in each country using an online dashboard to update the pandemic risk scores and contributions.

## Introduction

Since the declaration of the coronavirus disease 2019 (COVID-19) as a global pandemic by the World Health Organization (WHO) on 11 March 2020, there have been more than 24 million confirmed cases and more than 800,000 deaths worldwide as of 28 August 2020. Many countries have imposed preventive measures, such as city lockdown, travel restrictions, quarantine, widening social distancing, enhancing personal hygiene, etc, for stopping or slowing down further transmission of the disease. While continuing with the pandemic measures, it is also important to assess the pandemic risk for setting up preparedness plan and control measures for COVID-19^1^ especially for those countries with high risk of wide-spread transmission^2^.

A common pandemic risk assessment approach is by epidemiological modeling, where population is divided into at least three groups: susceptible, infected, and recovered^3–6^. This approach has been successful in understanding the disease transmission in a region through mathematical formulation of human interactions which may lead to infection, and the rate of recovery from infection. However, the susceptible-infection-recovery approach was designed fundamentally for closed populations^7^ and so restricted to regional studies. To supplement the epidemiological modeling, it is helpful to combine data from different regions or countries to study the pandemic situation, i.e. the status of the disease transmission in multiple regions or over the world, and to quantify the potential risk involved due to the COVID-19 outbreak. Two issues are of particular interest: prediction and control. Prediction means whether we can learn insights from data to produce early warning signals of pandemic and for preparedness^8–10^. Control refers to assessing the current pandemic risk severity for deciding proper measures during global pandemic^11,12^.

In this paper, we propose network analysis^13^ to aggregate data information from all over the world to construct dynamic pandemic networks for COVID-19. Network analysis has been applied to analyzing scientific collaborations^14^, to econometrics for assessing systemic risk and contagion effects in financial markets^15,16^, to medical research in studying gene co-expression, disease co-occurrence and global epidemics^17–20^, and to analyzing text^21^. We extend the network approach in the literature^9,22^ to study topological properties of COVID-19. There are three main features of our proposed network analysis. First, we make use of publicly available data, namely daily number of confirmed cases and daily accumulated number of infected people in each country to learn topological properties of dynamic pandemic networks and to visualize the propagation of COVID-19 for risk prediction and control. Second, we construct two pandemic risk scores and determine risk contributions from countries for pandemic prediction and control. The risk contributions of countries are helpful for setting preparedness plans and for assessing the severity of COVID-19 outbreak in specific regions. Finally, we compare the COVID-19 topological features with an independent network, the Erdos-Renyi model^23^, to understand the current pandemic situation and observe any signal for the COVID-19 outbreak to go away (for restarting economic activities).

In the healthcare management consideration, it is important, yet challenging, to assess pandemic risk of COVID-19 in the global perspective. In this study, we propose the network analysis and two pandemic risk scores using publicly available information on the number of confirmed COVID-19 cases, and the estimated number of currently infected people. We provide evidence that network statistics can yield early warning signals of the outbreak of COVID-19. The time series of the two pandemic risk scores and the respective risk contributions by countries can help to predict or ‘stress test’ the potential risk involved from releasing travel restriction measures between countries, and to estimate the severity of the pandemic for epidemic control. Simulations from a network assuming independent links between countries suggest that there is no sign for COVID-19 to go away in a few months. In summary, Figure 1 shows the work flow diagram of the pandemic risk assessment system developed in this paper. We started from publicly available confirmed cases, recovered cases and the number of deaths due to COVID-19, to constructing dynamic pandemic networks an1d their network statistics, and finally compiling the preparedness risk score (PRS), and the severity risk score (SRS) for risk assessment. Details of the work flow are given in Figure 1 and in *Methods*. For tracing the current pandemic risk of 164 countries, we have included the most updated risk scores and risk contributions in http://covid-19-dev.github.io/.

**Figure 1.**
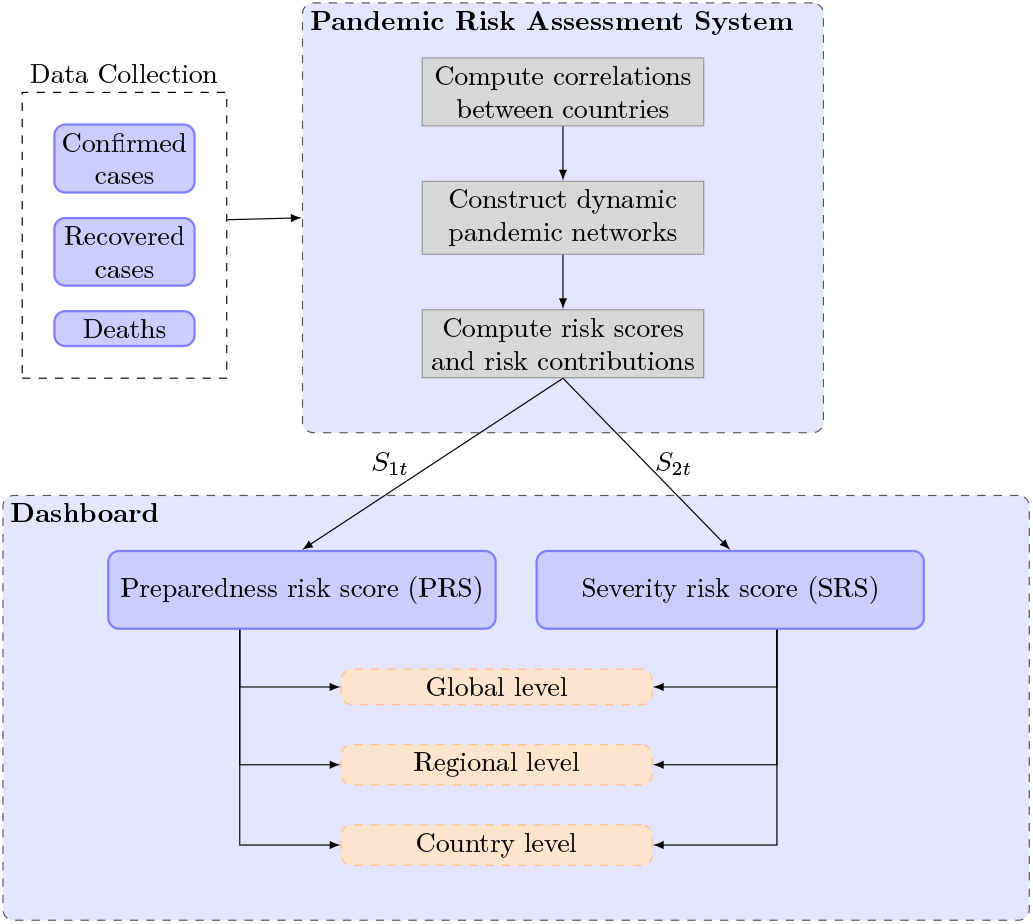
The pandemic risk assessment system work flow diagram.

## Results

### Network connectedness

As described in *Methods*, we followed the literature^9,22^ to construct time series of pandemic networks of 164 countries from February 2020 to mid August 2020. The countries were linked together in a particular day when the correlation of the increase in the number of confirmed cases in the past 14 days exceeds a certain level. This ‘co-movement’ of newly confirmed COVID-19 cases defines the links between countries and forms the pandemic networks. To quantify the network connectedness, we calculated the network statistics presented in *Methods*. Figure 2 shows the pandemic networks on 4 February 2020, 11 March 2020, and 11 April 2020. From the figure, we can visualize the pandemic networks are getting more dense from February to April 2020. This visual increase in connectedness was documented as an early warning signal of the COVID-19 pandemic in the literature^22^. The left panel of Figure 3 presents the number of edges, network density, clustering coefficient, and the assortativity coefficient of the COVID-19 pandemic networks from February 2020 to mid August 2020. The right panel of Figure 3 gives the corresponding network statistics for the an independent network, the Erdos-Renyi model, which are discussed in the *Discussion*. From the left panel of the figure, the actual number of edges in the pandemic network increases steadily from late February 2020 and rises sharply after the WHO’s declaration of the COVID-19 pandemic on 11 March 2020 to the peak on 18 March 2020. The sharp increase of the number of edges in the week of 12-18 March 2020 is partly attributed to having more countries reporting their confirmed COVID-19 cases, and partly due to more edges being formed because of the ‘common trend’ in the confirmed cases in more countries. The pattern of the number of edges in this particular week evidently confirms the pandemic announced by the WHO. The downward trend in the number of edges after 18 March 2020 signifies the time period of epidemic control of the further spreading for COVID-19 by countries which implemented various measures, like quarantine rules, travel restrictions, and enlarging social distancing. The downward trend also reveals the effect of those measures on the pandemic control and thus seeing less coherence in the occurrence of confirmed cases in the countries. The number of edges has been stable in April to June, though there seems to have a small tendency of rising up again in July.

**Figure 2.**
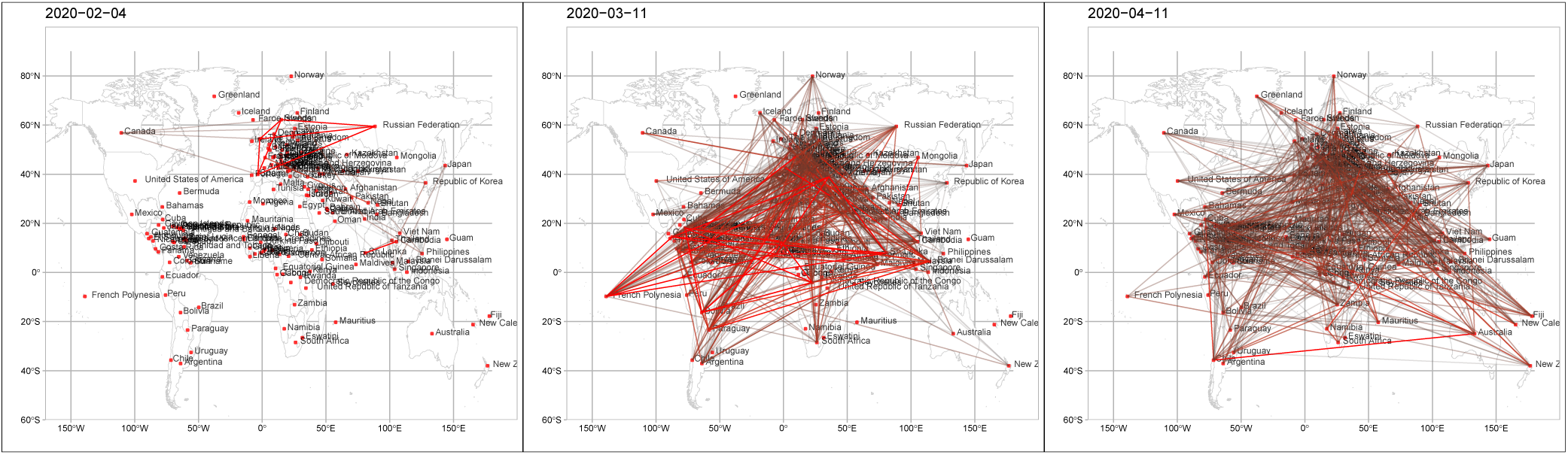
The COVID-19 pandemic networks on 4 February 2020, 11 March 2020, and 11 April 2020.

**Figure 3.**
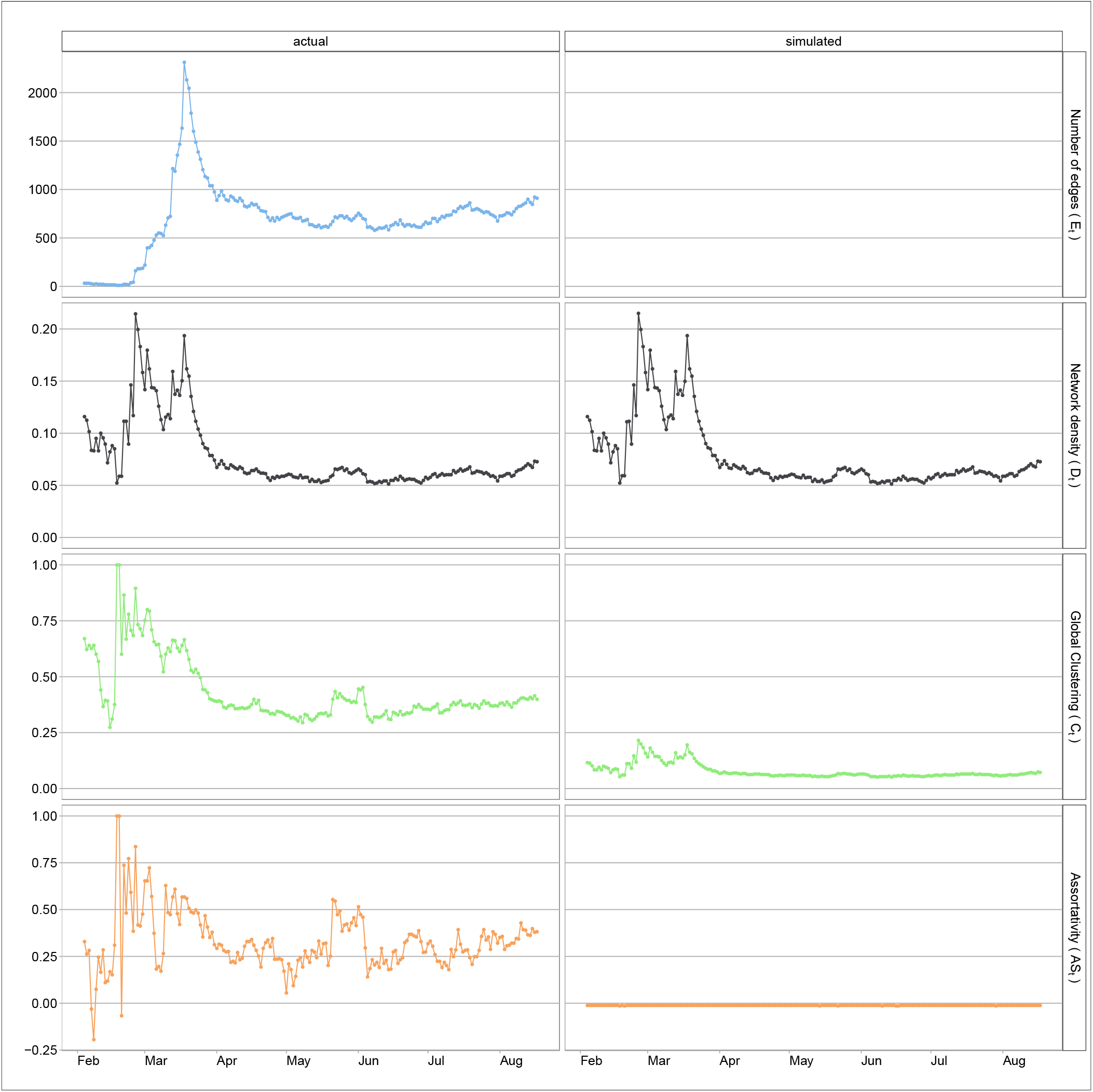
The time series plots of the network statistics, including the number of edges, network density, clustering coefficient and assortativity coefficient. The left column gives the network statistics for the COVID-19 pandemic networks, and the right column gives the corresponding statistics from an independent network, the Erdos-Renyi model (simulated networks)

The research in the literature^9^ proposed that we can use the network density, *D*_*t*_ in Figure 3, to generate early warning signals of the COVID-19 pandemic. We observe in the left panel of Figure 3 that *D*_*t*_ shows the first peak on 26 or 27 February 2020 which is analogous to ‘pre-earthquake phenomena’ as an alarm to a big earthquake some days later. The first peak of the network density, induced by the high connectedness of changes in confirmed COVID-19 cases from countries in late February, probably tells us that the pandemic was happening. There has been stronger evidence of co-occurrence of more confirmed cases in the countries, leading to a sharp increase in the network density in just two to three days in late February. There is another peak in the network density in mid March after the WHO’s declaration of the COVID-19 pandemic on 11 March 2020. Similar to the number of edges, the network density is quite stable in April to July.

The clustering coefficient at time *t* is a measure of how close the pandemic network at time *t* to a perfectly-linked network where all countries are linked together. In the extreme case (of the perfectly-linked network) where all countries’ confirmed cases increase simultaneously in the past 14 days, the countries will be well connected and can be viewed as one big pandemic region. Therefore, when the clustering coefficient is higher, the pandemic network will be more similar to the perfectly-linked situation, indicating stronger evidence of the global pandemic. From the clustering time series in the third row of the left panel in Figure 3, we observe that the highest value occurs on 26 February 2020, when we also observe the first peak in the network density. The clustering coefficients also present an early warning signal of the global COVID-19 pandemic. Unlike the network density, the clustering coefficients do not show another obvious peak in mid March. Although we still observe substantial confirmed COVID-19 cases in various countries, the clustering coefficients on around 18 March (when we see another peak in the network density) is smaller than that on 26 February 2020, implying that the pandemic network is less similar to the perfectly-linked network than what we see on 26 February 2020. The downward trend in the clustering coefficient probably implies that the global outbreak of COVID-19 was gradually controlled since early March, though it takes time for the actual confirmed cases to go down substantially. The clustering coefficient lies between 0.25 to 0.50 after March.

The assortativity coefficient, *AS*_*t*_ in equation (5), is to assess the assortative mixing of the vertices or countries in the pandemic networks. The last row in Figure 3 (in the left panel) presents *AS*_*t*_ from February 2020 to mid August 2020, where we see the value of *AS*_*t*_ to jump up to around 0.5 in late February. The substantial increase in *AS*_*t*_ in February implies that countries with a large number of connections tend to link with high-connection countries. This pattern of association of ‘similar’ countries (in terms of the degree *k*_*it*_ in equation (3)) can indicate possible inflection risk in social networks^24^. In the current COVID-19 pandemic situation, we can view this high *AS*_*t*_ as an indication of the disease outbreak since it is likely when a global pandemic occurs, high-connection countries tend to be linked together because they may have a simultaneous increase in the number of confirmed COVID-19 cases in a short period of time. It is not surprising to see that the time series pattern of *AS*_*t*_ is similar to that of *C*_*t*_ in the third row of Figure 3. The assortativity coefficient stays at around 0.25 after April 2020, except in late May to early June, it jumps up to around 0.5 for around two weeks.

### Pandemic risk sores

The preparedness risk score (PRS), *S*_1*t*_ and the severity risk score (SRS), *S*_2*t*_ are computed for *t* from 4 February 2020 to 17 August 2020. Both *ω*_*t*_ and *n*_*t*_ are measured in millions of people. We plot the standardized *S*_1*t*_, in which *S*_1*t*_ is divided by the total number of possible interactions (when all countries are directly linked and *ω*_*t*_ taken as the population sizes) in Figure 4 (left top panel). The first peak appears on 2 March 2020 when the standardized *S*_1*t*_ increases sharply from around zero to close to 0.1 in just a few days. Using 0.05 as a reference, i.e. 5% of the total interactions between people in the 164 countries, this sharp increase marks the first time when the risk score exceeds this reference, and can be regarded as an early warning signal of the COVID-19 pandemic. The second peak is found on 18 March 2020, a week after WHO’s declaration of COVID-19 as a global pandemic on 11 March 2020. This spike reaches a point close to 0.4, accounting for more than one-third of the total interactions between people in the 164 countries. After that, probably due to stringent measures imposed by various countries, including travel restrictions, community lockdown and enhancing social distancing, the standardized *S*_1*t*_ drops but quite slowly in mid April and May. The time series of the PRS stays mostly above 0.05 till mid June, indicating that the pandemic risk is still substantial even after three months of the WHO’s declaration with tremendous measures and efforts from various countries in preventing the transmission of COVID-19. What worries us is whether this standardized *S*_1*t*_ will go up again when some countries release travel restriction measures later. In fact, there is a tendency for *S*_1*t*_ to rise in July.

**Figure 4.**
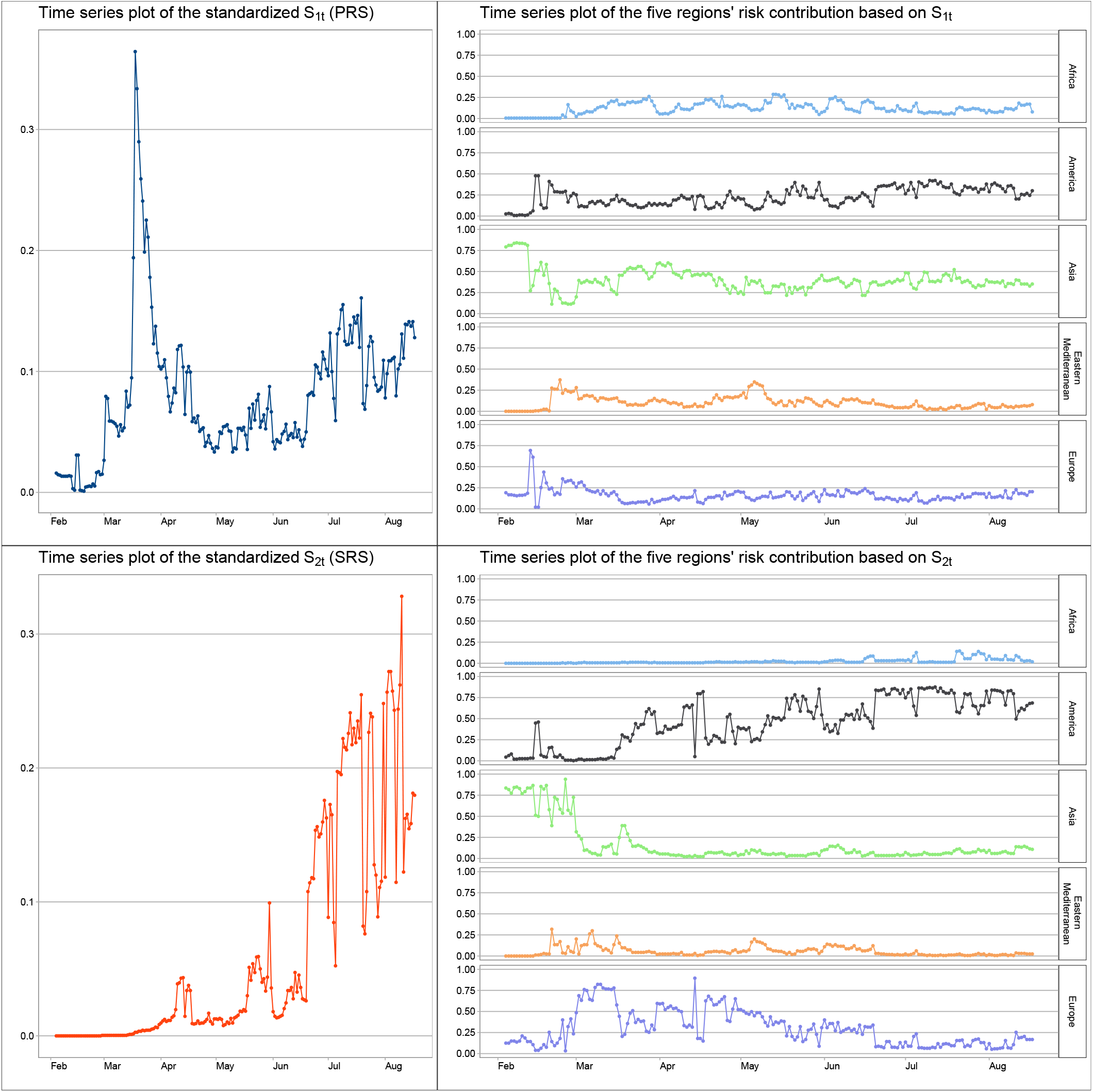
Plot of the standardized *S*_1*t*_ with the standardization done by all possible interactions between people in the 164 countries, and the standardized *S*_2*t*_ with the standardization done by all possible interactions between people in the 164 countries divided by 0.1%

Figure 4 (left bottom panel) displays the time series of the standardized *S*_2*t*_, in which *S*_2*t*_ is divided by the total number of possible interactions (which all countries are directly linked, with *ω*_*t*_ taken as the population sizes and *n*_*t*_ taken as 0.1% of the population sizes). The construction of *S*_2*t*_ is more sensitive to the confirmed COVID-19 cases and thus *S*_2*t*_ can serve as a measure of the COVID-19 outbreak severity. The first peak of *S*_2*t*_, signifying the first wave of the pandemic, is on 12-13 April 2020, roughly a month after the WHO’s declaration on 11 March 2020. There is improvement in reducing the number of confirmed cases and possibly also the pandemic network connectedness that we observe obvious drop in *S*_2*t*_ in late April and early May. However, another local peak is observed on 30 May 2020, which likely indicates the second wave of the outbreak in the second half of May. The recent increasing trend of *S*_2*t*_ appeared in June and July probably implies that the third wave of the outbreak of COVID-19 has come. The situation may get worse later if there is no preventive measure for further stopping the transmission of the disease in local community or if travel restrictions are going to be suspended or stopped. More worry is about the reluctance of wearing masks in hot weather in the summer.

### Risk contributions from countries

Figure 4 (right top pannel) gives the risk contributions of Africa, America, Asia, Eastern Mediterranean and Europe by grouping the *f*_*it*_, the risk contribution based on the first pandemic risk score *S*_1*t*_ in equation (8), in the five regions. We observe that Asia contributes most of the pandemic risk associated with *S*_1*t*_ in February 2020 and the Asia’s contribution stayed mostly between 0.25 and 0.5 after February. In the predictive perspective, reducing the mobility of the Asia’s population can be a reasonable preventive measure during the pandemic. Recently, there is an obvious increasing trend in the America’s contribution to close to 0.5 in July, indicating relatively higher risk contribution of transmission of the disease due to possible interaction between susceptible populations.

Figure 5 presents the heatmap of *f*_*it*_. Based on the construction of *S*_1*t*_ using mainly the interactions between two susceptible populations, this risk contribution is more on preventive preparation if there is chance for people from the two countries to interact. So the risk contribution *f*_*it*_ is like doing ‘stress testing’ of the pandemic risk. In early February, the risk score is mainly contributed by Asia, where the risk contributions from India and Philippines are around 0.3. From late February to early March, the risk score from non-Asian region, e.g. United States from America, Iran and Pakistan from Eastern Mediterranean, and Italy, Germany and France from Europe started showing significant contributions to the risk score. In particular, *f*_*it*_ of the United States has been quite high since mid February, signifying the early stage of the pandemic in the country. In late May and June, Ethiopia, Nigera from Africa, and Brazil from America also showed around 0.1 contribution that should be alerted.

**Figure 5.**
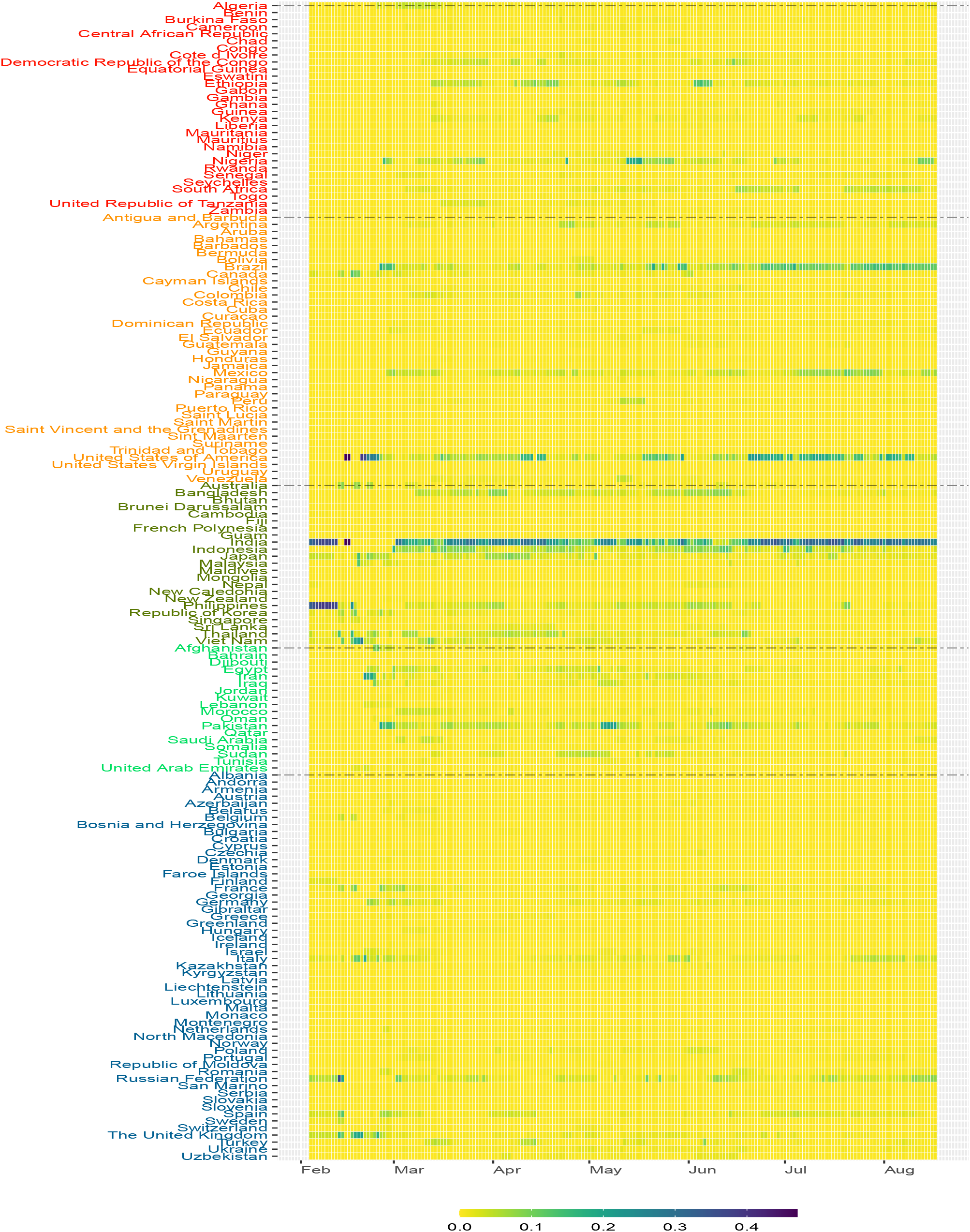
The heatmap of the risk contribution *f*_*it*_ based on the preparedness risk score *S*_1*t*_ for the 164 countries from February to mid August 2020

Figure 4 (right bottom pannel) gives the risk contributions of Africa, America, Asia, Eastern Mediterranean and Europe by grouping the *g*_*it*_, the risk contribution based on the second pandemic risk score *S*_2*t*_ in equation (9). Since *S*_2*t*_ is deduced from the number of possible interactions between people of a susceptible population and people of an infected population, the number of confirmed cases will play an important role in determining the risk contribution *g*_*it*_. From Figure 4, the pandemic risk based on *S*_2*t*_ was contributed mostly by Asia in February 2020, and mostly by Europe and America in March and April 2020. After April, the risk contribution from America climbed up to about 0.5, and exceeded 0.75 in July 2020. Figure 6 presents the heatmap of *g*_*it*_. This risk contribution reflects the severity of the COVID-19 pandemic risk from each country and thus is a viable measure for guiding us on the pandemic control of further transmission of the disease. In early February, the risk contributions of Japan, Philippines, Korea, Singapore and Thailand are quite high. The risk contributions are then dominated by Korea, Iran and Italy in early March, and followed by United States and Spain in late March. We can see that *f*_*it*_ of United States appears to be high in late February, earlier than seeing large value in *g*_*it*_, probably indicating that there was chance for United States to set up stringent control measures in late February before the widespread of the disease reflected in *g*_*it*_ afterward. On the contrary, India decided the national lockdown policy on 24 March 2020^25^. The lockdown policy may account for the fact that even though its *f*_*it*_ showing quite high value in February, its risk contribution indicated by *g*_*it*_ is very low in March and April. The cases of United States and India probably indicates the possible use of *f*_*it*_ as a measure for setting up timely stringent measures. In late April to June, the risk contribution *g*_*it*_ is dominated by Brazil, Italy, Peru, United Kingdom, United States and Russia. India and South Africa’s risk contributions have increased in July. The severity of the COVID-19 pandemic risk in those countries with relatively high *g*_*it*_ cannot be ignored.

**Figure 6.**
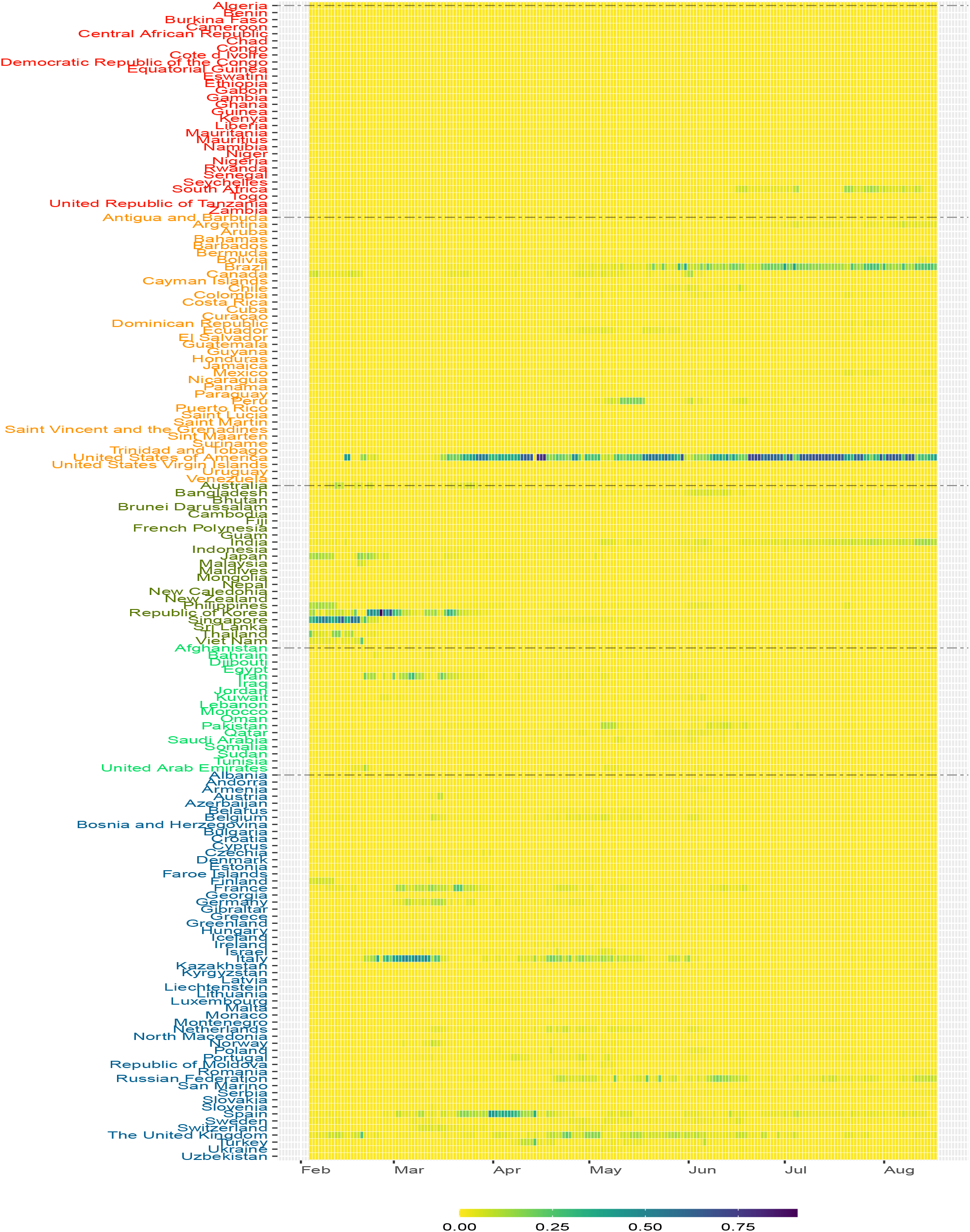
The heatmap of the risk contribution *g*_*it*_ based on the severity risk score *S*_2*t*_ for the 164 countries from February to mid August 2020

### Discussion

To assess the current pandemic status of COVID-19, we simulate networks from the Erdos-Renyi model^23^. Specifically, at each time *t*, we generate 10,000 networks with edges independent of each other. To maintain the same level of network density, two vertices in a simulated network are linked together with the probability *D*_*t*_. The right panel of Figure 3 displays the time series of the average of the network statistics from the 10,000 simulated networks. The outbreak of a disease likely makes the pandemic network to follow some structures which deviate from the the Erdos-Renyi model. For example, in the COVID-19 pandemic, the common trend in the confirmed COVID-19 cases observed between countries *i* and *j* and countries *j* and *k* may imply a relatively higher chance to observe a common trend on the COVID-19 confirmed cases in countries *i* and *k* as well (this is the transitivity property in social networks). Therefore, by comparing the discrepancy between the time series patterns of *C*_*t*_ and *AS*_*t*_ between the COVID-19 pandemic networks and the Erdos-Renyi model, we can find hints as to when the global COVID-19 pandemic will go away. From Figure 3, the clustering coefficients of the pandemic networks are well above that of the the Erdos-Renyi model with the same network density *D*_*t*_. Although there is sign for the assortativity coefficients of the pandemic networks to approach zero, the assortativitiy coefficient of the Erdos-Renyi model, there is no strong evidence that COVID-19 will go away shortly after August 2020.

The preparedness risk score *S*_1*t*_ accounts for the possible interactions between susceptible population of countries being linked together in the pandemic networks, and can be used to quantify the risk of transmission of the COVID-19 between countries. The possible transmission risk is attributed to substantial percentages of asymptomatic transmission or presymptomatic transmission^26^. Therefore, this *S*_1*t*_ score is a kind of ‘stress testing’ measure^27^ which evaluates the potential transmission risk involved if travel restriction policies are canceled or become less strict. From the time series plot of the standardized *S*_1*t*_ in Figure 4, the transmission risk, or *S*_1*t*_, increases sharply on 2 March 2020 to 0.08, and 18 March 2020 to 0.36, implying more than 1/3 of all possible interaction between susceptible populations of the 164 countries. The first peak on 2 March 2020 (well before the WHO’s declaration on 11 March 2020) can be regarded as an early warning signal of the COVID-19 global pandemic. The second peak on 18 March 2020 marks the rapid spreading of the disease in March and early April, when the number of cases of Italy, Spain, France and other European countries increase substantially^3,28,29^. After reaching the peak on 18 March 2020, the transmission risk went down in April and May, probably because of a series of travel restrictions and other lockdown measures which significantly stopped the mobility of susceptible population between countries to almost completely disappeared. However, *S*_1*t*_ didn’t go down further and stayed at around 0.05 in April and May, implying that the transmission risk had not been reduced to a level that would enable the release of travel restriction measures. Even worse is that the increase in the transmission risk was reignited in late June and July.

The risk contribution *f*_*it*_ in equation (8) based on *S*_1*t*_ quantifies the relative impact of country *i* at time *t* in transmitting COVID-19 through interaction between susceptible populations. The higher the value of *f*_*it*_, the higher is the potential impact from country *i* at time *t* with respect to the transmission risk. The risk contribution *f*_*it*_ can be interpreted as a relative transmission risk measure which can be considered by governments to set up preventive measures by revisiting their epidemic control policies applied to different regions from time to time. For example, we observe that *f*_*it*_ of Nigeria and South Africa (of Africa), Brazil, Mexico and United States (of America), India and Indonesia (of Asia), Pakistan (of Eastern Mediterranean) and Russia and Spain (of Europe) are relatively high in June and July. Governments are recommended to be more alerted when revisiting any preventive measure with those countries whose *f*_*it*_’s are high.

The severity risk score *S*_2*t*_ calculates the possible interactions between susceptible populations and the currently infected population of countries being linked together in the pandemic networks. The score reflects the current severity of the COVID-19 outbreak due to the transmission of disease from infected people who may have symptoms or due to presymptomatic transmission. The evolution of pandemic risk severity reflected by the time series pattern of *S*_2*t*_ provides a reference for governments to revisit their current epidemic control measures to see if it is necessary to revise or to strengthen the current measures for preventing their medical systems from overloading too much or breaking down. In Figure 4, we observe from *S*_2*t*_ the first wave of COVID-19 outbreak in early to mid April when there is substantial increase in the confirmed COVID-19 cases in Europe^28^. There is an obvious drop in *S*_2*t*_ in late April till mid May, probably due to the lockdown control in European countries^30^. The second wave of the COVID-19 outbreak fell in late May when there were graduate economic reactivation activities implemented. Another sharp increase in the risk score started in late June and the increasing trend continued in July and August, signifying the third wave of the outbreak. The high value in *S*_2*t*_ can give an alerting signal to governments about the need to slow down the recovering of social activities or the revival of economy.

Although *g*_*it*_ refers to the risk contribution due to interactions between infected people and susceptible population of countries, high *g*_*it*_ also implies high risk of ‘domestic transmission’ if people from different regions of a country can have physical contact, for example, through school resumption, social, or religious activities. Before and during the first wave of COVID1-19 outbreak in March and April, we see relatively high *g*_*it*_ in Figure 6 in France, Germany, Italy, Spain, United Kingdom and United States. After imposing strict countrywise lockdown policies, the risk contributions of many European countries, except Russia, went down in May. In terms of *g*_*it*_, the pandemic risk severity of United States does not significantly lower in May. We also see the transmission of the disease to South America where the risk contribution of Peru is quite high in May. In July, the total risk contribution in Brazil and United States accounts for around 80% of the overall *S*_2*t*_ and we also see substantial increase in risk contribution in South Africa. Governments and healthcare specialists may not ignore relatively high *g*_*it*_ which may indicate the relative severity of COVID-19 pandemic risk in countries, especially if those countries are financial, economic or even travel hubs in the region.

For tracing the latest COVID-19 pandemic status, we have included the most updated risk scores and risk contributions in http://covid-19-dev.github.io/.

## Methods

As presented in Figure 1, the first step of the proposed pandemic risk assessment system is to construct dynamic pandemic networks using publicly available confirmed COVID-19 cases.

### Network statistics

Let *X*_*i,t*_ be the number of confirmed COVID-19 cases of country *i* in day *t*. Following the literature^9,22^, we obtain the daily changes for each country as 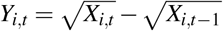. To construct dynamic pandemic networks, we calculate *ρ*_*i j,t*_, the correlation between country *i* and country *j*’s daily changes in day *t* using the observations (*Y*_*i,t*−*k*_,*Y*_*j,t*−*k*_), for *k* = 0, …, 13. After that, we construct the pandemic network at time *t* by defining *A*_*i j,t*_, which is the (*i, j*)th element of an adjacency matrix of the pandemic network at time *t*:

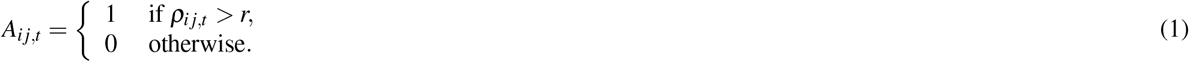

In other words, an edge/link between country *i* and country *j* is formed in the pandemic network at time *t* if *ρ*_*i j,t*_ > *r*. Since the earliest report of the number of confirmed cases by the WHO in late January 2020, we have more countries in giving us the data of *X*_*i,t*_. To study the statistical properties of the pandemic network from February to mid August 2020, we start from recording the number of countries having confirmed cases records at time *t*, denoted by *V*_*t*_, which is less than or equal to 164 (country names listed in Figures 5 and 6). Statistically, *V*_*t*_ is the number of vertices of the pandemic network at time *t*. When a country has zero *Y*_*i,s*_ in 14 consecutive days (*s* = *t* −13,…, *t*), it will be excluded from *V*_*t*_. Figure 2 shows the pandemic networks at *t* = 1 (4 February 2020), *t* = 36 (11 March 2020) and *t* = 67 (11 April 2020), from which we observe increasing level of connectedness in the pandemic networks from early February to early April^22^. To quantify the network connectedness and to summarize other network properties, we consider four network statistics at time *t, E*_*t*_ (the number of edges), *D*_*t*_ (the edge density), *C*_*t*_ (global clustering coefficient), and *AS*_*t*_ (assortativity). Since we define the edge of the pandemic network by the event that the correlation *ρ*_*i j,t*_ exceeds the threshold *r*, the number of edges, *E*_*t*_, gives the number of pairs of countries showing a ‘common trend’ in terms of the confirmed COVID-19 cases in the past 14 days, including time *t*. In this paper, we follow the literature^9^ to set *r* = 0.5. The network density, *D*_*t*_, defined by

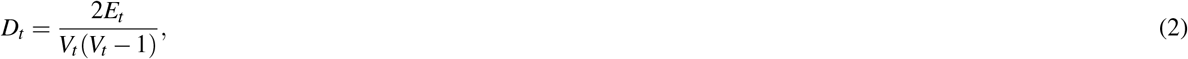

is to understand how dense the pandemic network is at time *t*. The literature^9^ presents visual evidence on using *E*_*t*_ to give an early warning signal of the COVID-19 pandemic.

To capture how strong vertices tend to be linked together, we calculate a clustering coefficient of the pandemic network at time *t*. For vertex *i*, define its local clustering coefficient at time *t* as

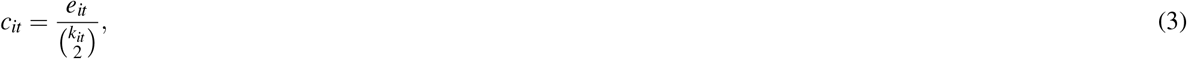

where *e*_*it*_ is the number of connected pairs among the neighbors of vertex *i* at time *t*, and *k*_*it*_ is the number of neighbors (or degree) of vertex *i* at time *t*. The numerator in equation (3), *e*_*it*_, also counts the number of triangles formed by vertex *i* in the pandemic network at time *t*, and 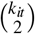 is the number of connected triples (a subgraph of three vertices connected by two edges) at time *t* connecting vertex *i*. The *c*_*it*_ measures the tendency for vertex *i* to form triangles with its neighbors. The clustering coefficient, denoted by *C*_*t*_, is a weighted average:

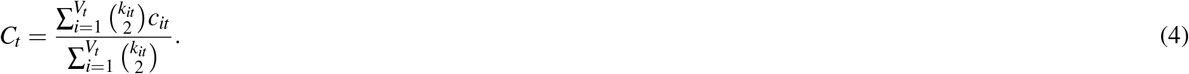

The clustering coefficient *C*_*t*_ measures how strong vertices/countries in the pandemic network at time *t* are bounded together in clusters. In the extreme case where *C*_*t*_ = 1, all pairs vertices/countries are connected by edges in the pandemic network at time *t*. In other words, the higher the *C*_*t*_, the stronger the evidence of global pandemic is revealed in the pandemic network.

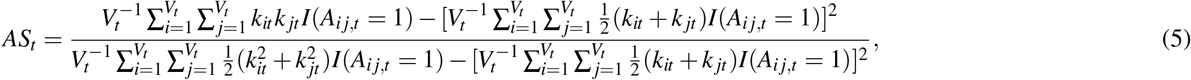

In addition to the network density and the clustering, we also consider the assortativity of networks^31^, which is used to describe assortative mixing properties in networks. Specifically, we evaluate the tendency for countries with similar degrees to link together in the pandemic networks. The degree of country *i* is defined as the number of countries connecting that country at time *t* and is given by *k*_*it*_. Using the network terminology, *k*_*it*_ is the number of edges incident to vertex/country *i*. In epidemiological perspectives, the risk of individuals’ infection may be related to the assortativity (high degree vertices tend to link with other vertices with high degrees) in social networks^24^. In this paper, we apply this assortative mixing concept to the pandemic networks constructed to understand how the assortative mixing properties change over time during the COVID-19 pandemic. Statistically speaking, we calculate an assortativity coefficient which measures the correlation of the degress of the vertices in the pandemic networks. At time *t*, the assortativity coefficient is defined by equation (5) where *I*(*E*) is an indicator function whose value is equal to one if the event *E* is true and zero otherwise. We attempt to learn any insight from this assortativity coefficient on possible early warning signals of a global pandemic like COVID-19.

### Pandemic risk scores

In this paper, we propose two pandemic risk scores defined by the dynamic pandemic networks represented by *A*_*i j,t*_ in equation (1), the adjacency matrix at time *t*. Specifically, we aggregate the link information in *A*_*i j,t*_, susceptible population sizes (or population at risk) of each country, and the number of confirmed cases, *X*_*i,t*_, to construct two scores which help us to assess the potential transmission risk across countries, and the severity of the global pandemic risk due to COVID-19. Define the first risk score as the preparedness risk score (PRS)

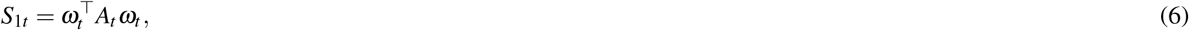

where *ω*_*t*_ is the vector of population size of each country (based on the World Bank population figures in 2018) subtracted by the total number of confirmed cases in each country up to time *t*. In other words, the *i*th element of *ω*_*t*_, denoted by *ω*_*it*_, represents the population of country *i* at risk or susceptible at time *t*. Since 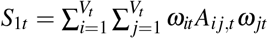 in equation (6) counts the total number of possible interactions of susceptible population contributed from all pairs of countries which are linked together at time *t*. This PRS accounts for the risk of asymptomatic transmission or presymptomatic transmission^32^ due to possible interaction between people in the two countries. These two kinds of transmission are usually hidden in the population and is useful to quantify the pandemic risk especially when the transmission between two countries cannot be completely stopped by travel restrictions or other infection prevention policies. Therefore, the risk score, *S*_1*t*_, can help us to project the severity of the COVID-19 outbreak for preparedness if existing travel restrictions or lockdown schemes are released.

We define the second risk score as the severity risk score (SRS)

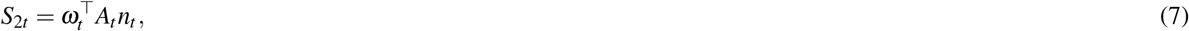

where *n*_*t*_ is the vector of the total number of currently infected people at time *t*. In other words, the *i*th element of *n*_*t*_, denoted by *n*_*it*_ is calculated as 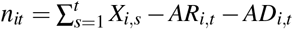, where *AR*_*i,t*_ and *AD*_*i,t*_ are the accumulated number of recovered cases and accumulated number of deaths due to COVID-19 in country *i* at time *t*. It is easy to see that 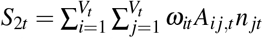. The rationale behind *S*_2*t*_ is to count the number of possible interactions between currently infected cases of one country and all people at risk in another country, if the two countries are linked together. The product of *ω*_*it*_*n* _*jt*_ is similar to the classical SIR model^7^, where the number of susceptible people at time *t* multiplied with the number of infected people at time *t* is used to determine the rate of change of the number of susceptible people. This SRS accounts for presymptomatic transmission due to possible interaction between susceptible population in one country and currently infected cases in another country before the confirmed cases are identified and forced-isolated or quarantined. The SRS, *S*_2*t*_, can help assess the severity of the COVID-19 due to the current level of infections and plan for outbreak control measures. We present this step in computing the PRS and SRS in the work flow diagram in Figure 1.

As in the literature^33^, we can define the risk contribution by country *i* on the PRS as

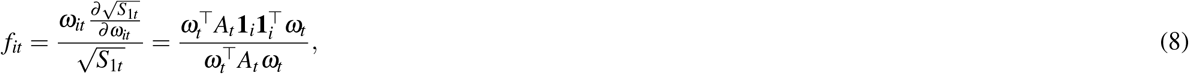

where **1**_*i*_ is a column vector with the *i*-th entry equal to 1 and 0 otherwise. Using the Euler rule, we can show that ∑_*i*_ *f*_*it*_ = 1. From equation (8), *f*_*it*_ is approximately equal to the ratio of the percentage change in 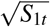 over the percentage change in *ω*_*it*_. If a particular country *i* has high *f*_*it*_, reducing the mobility of the people (at risk) in that country may lead to substantial reduction in *S*_1*t*_, thereby lowering the global pandemic risk. Similarly, we can define the risk contribution by country *i* on the SRS as

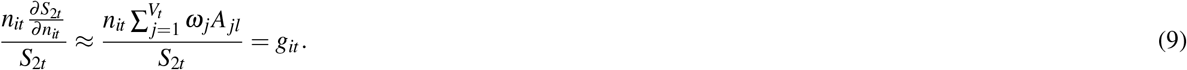

Again, we can show that ∑_*i*_ *g*_*it*_ = 1. Equation (9) implies that *g*_*it*_ is approximately equal to the ratio of the percentage change in *S*_2*t*_ over the percentage change in *n*_*it*_. If a particular country *i* has high *g*_*it*_, quarantining currently infected cases and enhancing social distancing measures as best as we can help ‘block’ the interaction of infected people with people at risk, and may lead to substantial reduction in *S*_2*t*_, thereby achieving better control on the global pandemic risk. In summary, for outbreak prediction or prevention, *S*_1*t*_ is recommended for policymakers to make reference to. For outbreak control, *S*_2*t*_ is more meaningful.

The risk contributions *f*_*it*_ and *g*_*it*_ with respect to the PRS and SRS can be computed on the global, regional, and country levels (see Figure 1). For active pandemic risk monitoring, the most updated PRS, SRS, *f*_*it*_, and *g*_*it*_ can be accessed in http://covid-19-dev.github.io/.

## Data Availability

The data are available in in http://covid-19-dev.github.io/.

## Data availability

The authors confirm that the real data sets used in this study are freely available to public at the time of writing by accessing the webpages: https://github.com/CSSEGISandData/COVID-19 and https://www.who.int/emergencies/diseases/novel-coronavirus-2019/situation-reports (accessed on 19 August 2020).

## Acknowledgements

The work described in this paper was partially supported by a grant from the Research Grants Council of the Hong Kong Special Administrative Region, China (16307217).

## Author contributions

So, M.K.P. and Chu, A.M.Y. conceptualized the study. So, M.K.P. and Chan, J.N.L., collected and analyzed the data. So, M.K.P., Chu, A.M.Y. and Tiwari, A. interpreted the results. So, M.K.P. and Chu, A.M.Y. drafted the manuscript. So, M.K.P., Chu, A.M.Y. and Tiwari, A. finalized the manuscript. All authors reviewed the manuscript.

## Competing interests

The authors declare no competing interests.

## Notes

### Competing Interest Statement

The authors have declared no competing interest.

